# Clinicians’ Rationale for Editing Ambient AI–Drafted Clinical Notes: Persistent Challenges and Implications for Improvement

**DOI:** 10.64898/2026.02.20.26346729

**Authors:** Yawen Guo, Di Hu, Ziqi, Emilie Chow, Steven Tam, Danielle Perret, Deepti Pandita, Kai Zheng

## Abstract

**Objective:** The use of ambient AI documentation tools is rapidly growing in US hospitals and clinics. Such tools generate the first draft of clinical notes from scribed patient-provider conversations, which clinicians can then review and edit before signing into electronic health records (EHR). Understanding how and why clinicians make modifications to AI-generated drafts is critical to improving AI design and clinical efficiency, yet it has been under-studied. This study aims to address this gap.

**Materials and Methods:** We conducted semistructured interviews with 30 clinicians from the University of California, Irvine Health who used a commercial ambient AI tool in routine outpatient care. We invited them to describe how and why they edited AI drafts based on both their personal experience and review of some real-world examples identified from our previous studies.

**Results:** Modifications to AI drafts were primarily made to improve clinical accuracy and specialty-specific precision, reduce medico-legal and liability risk, and meet billing, coding, and documentation standards. Such editing was necessary due to reasons such as transcription errors, speaker attribution mistakes, overconfident statements without evidence, missing key clinical details, and AI’s lack of information about the patient context.

**Conclusion and Discussion:** Improving ambient AI documentation will require coordinated effort from vendors, institutions, and clinicians. Key targets include core model reliability (e.g., transcription accuracy), specialty-and encounter-level customization, clinician-level personalization, more effective EHR integration, and institutional support (e.g., training, governance, and standardized review guidance), complemented by clinicians’ adaptive communication strategies that strengthen human–AI collaboration.

## Background and Significance

Ambient artificial intelligence (AI) documentation tools are rapidly becoming one of the most widely adopted generative AI tools in clinical care, which use visit audio to draft clinical notes and aim to reduce documentation burden and clinician burnout. [1–4] Early evaluations suggest adoption may improve documentation efficiency and productivity, supporting health enterprise growth. [5–8] Many clinicians report positive experiences, such as feeling more present with patients and reducing after-hours documentation. [6,9] At the same time, emerging concerns highlight the need for additional investigation for ambient AI performance and how it impacts care in routine clinical settings. Clinicians report persistent challenges, including: lengthy or underspecified note sections, problems with accuracy, with barriers in non-English encounters. [9–11] Importantly, the central role of clinical documentation in medico-legal accountability has raised concerns about downstream effects on billing, including potential upcoding, higher spending, and productivity pressures that may undermine clinician well-being. [12] Continued ethical and legal questions remain, including consent and transparency for patients and staff, and uncertainty about liability and retention when AI is used in clinical care. [13]

Prior work analyzing ambient AI use demonstrates that clinicians frequently edit AI drafts and that editing varies by note section, specialty, and individual documentation style. [14] Content analyses have also described recurring edit patterns, such as correcting factual errors, adding specialty-specific details, and reorganizing content for clarity. [15] However, these studies mainly characterize what changed in the note and do not fully capture why clinicians conduct specific edits or what persistent challenges are shaping those decisions. Because editing burden directly mediates the efficiency gains promised by ambient AI, understanding the drivers of edits is essential to realizing both clinician-experience and financial return-on-investment benefits.

We conducted semi-structured interviews with clinicians who used ambient AI tools in routine care and applied thematic analysis to examine (1) clinicians’ intent behind common edits, (2) and persistent challenges that shape editing decisions and trust. By linking clinicians’ explanations to recurring edit patterns observed in our prior studies [14,15], our findings identify practical targets for improving ambient AI note quality, workflow integration, and governance.

## Method

### Participant recruitment and interview procedures

We recruited clinicians from the University of California, Irvine Health (UCI Health), where a commercial ambient AI documentation tool Abridge (Abridge AI, Inc., Pittsburgh, PA, USA) was implemented in outpatient settings across multiple provider roles. Using an internal e-mail list of active users, we identified 87 clinicians with over 50 ambient AI–assisted encounters and 14 informatics clinician champions who used ambient AI and were extensively engaged in medical informatics research. We invited eligible clinicians via e-mail and interviewed 30 participants between January to February 2026; recruitment stopped when we judged thematic saturation had been achieved. Interviews were conducted remotely via Zoom and each lasted approximately 45 minutes. Participants provided verbal consent for audio recording. Recordings were stored in the Zoom cloud and transcripts were generated via Zoom automated transcription with only obvious transcription errors corrected. Transcripts were de-identified prior to coding, and access to the dataset was restricted to study authors. Participants received a $50 Starbucks gift card after completing the interview.

We used a semi-structured interview method to ensure consistent coverage of core topics while allowing participants to expand on experiences they considered relevant. In developing the interview guide, we drew on our prior content analysis of ambient AI draft–final note pairs, which identified five common edit patterns, including factual corrections, specialty-specific refinement, diagnostic certainty calibration, professionalization of transcript-like text, and restructuring and condensing. [14,15] These patterns informed an a priori framework that guided both interview domains and initial coding. We presented standardized, de-identified editing examples which aligned with these edit patterns and asked clinicians to interpret the edits and explain what drove their decisions. The interview protocol and example edits are provided in Supplementary Material S1. This study was reviewed and approved by the University of California, Irvine Institutional Review Board (UCI STUDY00000824-IRB).

### Qualitative coding and data analysis

We conducted thematic analysis to characterize clinicians’ perspectives, starting from the a priori framework described above and iteratively refining codes as new concepts emerged. Analyses were conducted in Atlas.ti (version 9.24.3; ATLAS.ti Scientific Software Development GmbH, Berlin, Germany). Three authors (YG, DH, ZY) jointly developed and refined the codebook through iterative rounds of double-coding (two interviews per round) and consensus discussions to resolve discrepancies and clarify definitions. One author (YG) then applied the refined codebook across the full dataset and led synthesis, supported by analytic memos and an audit trail. Within the a priori domains, we identified inductive subthemes by examining code prevalence, co-occurrence, and variation across interviews. To strengthen interpretive validity, we conducted member checking with two interview participants (EC, ST) who reviewed summary interpretations of key findings.

## Results

### Study Overview

A total of 30 clinicians participated in the interview study, yielding an overall response rate of 29.7%. Participants included 15 primary care clinicians and 15 subspecialists. Most were attending physicians (n=27); the remainder were nurse practitioners (n=2) and a physician assistant (n=1). Subspecialties represented included rheumatology (n=3), neurology (n=2), obstetrics and gynecology (n=2), neurosurgery, oncology, colon and rectal surgery, gastroenterology, integrative medicine, ophthalmology, radiation oncology, and hematology/oncology (n=1 each).

We analyzed interview transcripts, guided by findings from our prior note-pair content analysis, with a focus on emerging themes related to (1) clinicians’ revision intents, (2) the precipitating challenges that limited AI effectiveness, and (3) recommendations for improvement. In the following sections, we summarize the key intents driving edits and the recurring challenges clinicians described as barriers to achieving those intents. We then present clinicians’ recommendations for tool and implementation improvements.

### Clinician Editing Intents for Ambient AI Documentation

#### Correct factual, transcription, and attribution errors to protect record accuracy

Clinicians edit ambient AI drafts to prevent inaccuracies. Several clinicians emphasized that transcription mistakes were clinically consequential, especially when small lexical differences changed meanings, such as misspellings of “less common medications” including infusion agents used for chemotherapy (P17, P19, P20, P26). They linked terminology use errors to real-world speech conditions, including accent or unclear audio (P1, P11, P13, P17, P20–22, P27). One clinician summarized this limitation as it “can’t get it exactly” (P21). Additionally, identity or speaker attribution issues occurred, such as misspelled names, incorrect pronouns, or content assigned to the wrong speaker. When family members, partners, staff, or interpreters were present, clinicians reported the draft could carry over non-patient discussion, include symptoms described by someone other than the patient, or mismatch speakers with content(P1, P10, P12, P14, P21, P27). In some cases, misattribution led to clinically incorrect history, such as assigning a spouse’s transplant status to the patient, or pulling in a chemotherapy-order discussion that occurred when a nurse entered the room to update messages for another patient (P5, P10, P15, P18, P29), so clinicians needed to correct the inaccuracies manually.

#### Achieving specialty-specific precision and completeness in drafted notes

Clinicians described editing ambient AI drafts to make them more specialty-specific and complete, particularly when drafts “sounded” generic and did not reflect specialty priorities or clinicians’ differential reasoning (P14–16, P17–19, P22–23, P25, P27–28, P30).

Subspecialists emphasized that drafts often lacked differential framing and appropriate prioritization, often requiring rewriting note content to match how they practice (P16–17, P19, P23, P25, P27–28, P30). One participant summarized the mismatch as the AI drafts “ sounded like an internal medicine doctor… I’m a subspecialist” (P16). A psychiatry participant described the A&P drafts as “severely lacking” because their work involves “much more nuance” including “reading between the lines and inferring what patients are saying versus what they may mean” (P17). Rheumatology clinicians noted that important specialty constructs were often under-captured, such as documenting “low, moderate, or high disease activity”. They also reported that counseling was often summarized only as a vague mention of “risks,” rather than documenting specific risk-benefit discussions, side effects, or consent-related details (P15, P19, P24).

Beyond specialty-appropriate framing, clinicians also edited drafts to address omissions of clinically salient information. For completeness, participants noted that edits were often triggered by omissions of clinical details that the tool failed to capture, including missed review-of-systems questions, inconsistent capture of advance care planning and Medicare Annual Wellness functional/social assessments, and omission of negative screening histories unless a positive finding was present (P7–9, P13–14, P19, P21, P26, P30). Several described adding pertinent negatives (e.g., denied symptoms or cancer history) that they reported eliciting during the visit but that were not captured in the draft, to preserve clinical reasoning; as one clinician put it, “If I’m editing the history, I’m usually adding pertinent negatives that I feel the AI did not catch” (P11).

Participants emphasized that clinical specificity is not simply adding more detail, but preserving distinctions that affect interpretation and downstream management. They described these distinctions as involving symptom characterization, chronology, and diagnostic labeling, which were sometimes omitted or blurred in AI drafts (P22, P27–28). For example, one clinician noted the draft might say “fractured wrist” without “specifying where exactly, or the exact type of surgery,” reducing usefulness for follow-up care (P22). Clinicians noted that the draft could compress step-by-step symptom narratives into vague summaries and might retain an earlier version of the patient story unless later clarifications were explicitly restated for the record (P27, P28).

Clinicians also described restructuring content that the draft placed in the wrong note section or attributed to the wrong diagnosis. Several noted blurred boundaries between subjective report and clinician interpretation, with labs, imaging, or assessment statements appearing in the HPI (P4, P5, P13, P17, P20). One clinician explained that the AI draft mis-presented the clinician diagnosis “as if the patient said it” (P13). Others reported related sectioning issues, including placing patient-reported results or diagnoses into Results or A&P without preserving them as history (P7), or failing to integrate later-elicited history back into the HPI (P11). At the assessment level, clinicians said the draft could “latch[ed] on” to secondary issues and turn keywords into standalone diagnoses (P6, P9, P29), sometimes splitting a single condition into multiple diagnoses (P9) or adding historical diagnoses mentioned during the visit to A&P (P29).

#### Coding requirements, medico-legal safeguards, and evidence-aligned certainty

Clinicians edited ambient AI drafts to ensure billing, compliance, and medico-legal requirements were met, particularly in the A&P where coding depends on diagnostic specificity, documented decision-making, and severity-of-illness detail (P1, P5, P13, P25, P28). Several noted that the tool might suggest a diagnosis but still “doesn’t fill out the billing form” for level-of-service requirements, prompting them to add severity considerations and ensure documentation supported billing and risk-adjustment expectations (P1, P13, P25). Some explicitly linked these edits to documenting reasoning, describing it as important “from billing medico-legal standpoints” (P2, P28). When key elements were missing, participants described targeted additions, including details reflecting time spent, counseling, and options discussed, and relevant screening or history elements discussed during the encounter (P5, P7–9, P18–19, P28).

For example, oncology clinicians highlighted that consent documentation often requires substantially more specificity for billing than what is naturally said during patient-facing counseling: “for billing purposes, they want us to put in all the detail… list everything, just to be able to bill” (P5). Several clinicians further emphasized that time-based billing hinges on capturing not only actions taken but also counseling and options considered: “talking about things that are not done is still time we spent and options we offered… it justifies billing,” and “the more detail we have in the note, the more coding and billing we can do to justify the level of the visit” (P18). Additionally, a neurosurgeon noted that documentation sometimes must “satisfy several criteria before the insurance company would approve an operation,” and described ensuring the history clearly captured “a certain number of weeks of physical therapy or conservative measures… [and] prior trial of conservative measures” when the draft was not clear (P3).

Clinicians also described routine edits to align certainty with evidence and to reduce liability and patient confusion (P1–2, P4, P6, P11, P14–16, P17–19, P22, P24, P26–27, P29-30). They emphasized that documentation should reflect what is known versus still being evaluated, particularly early in a workup (P1, P7–9, P14–16, P18–19, P22, P24, P26–27). Several felt the draft could flatten probabilistic reasoning into definitive statements, for example, “I say ‘possibly this’… and the AI writes it as if it’s definitive” (P19), or “if I don’t explicitly say ‘these tests are to confirm,’ then it writes the diagnosis as confirmed” (P18). To restore evidentiary alignment, clinicians inserted qualifiers and conditional plans and avoided prematurely closing the diagnostic narrative, including when unconfirmed problems appeared in the A&P or when an intended hedge such as “likely a disc herniation” was missing (P1, P8, P14–16, P15, P27). They also described hedging as a communication and risk-management practice, such as using “low suspicion” as “medico-legal protection” (P2, P24).

#### Edit for professional, standardized notes and efficient clinical review

Across interviews, clinicians described editing ambient AI drafts to meet professional documentation standards so notes read like clinician-authored records rather than verbatim narratives (P2, P8, P9, P11–15, P17, P21, P22, P24, P25, P27–29). For example, participants wanted notes that “look professional when somebody reads it” (P14) and reflected how they were taught to write, including that it “shouldn’t read like a transcript” (P21). Edits often focused on avoiding informal wording and colloquialisms, and standardizing terminology, including converting spoken exam language into “medical terminology” (P4, P9, P11, P13, P17, P21, P23, P24, P27, P29). Some preferred more understandable and actionable language for the after-visit summary when the note was facing patients who may have limited health literacy (P29).

In addition, AI drafts were often “too wordy and paragraphy” (P2) and included redundancy, low-yield detail, or repeated plans, so clinicians condensed content, trimmed small expansions that accumulated, and shortened contents they felt were “too long” for other providers or patients to read (P5, P7, P9, P13–15, P21, P23, P24).

Workflow and EHR integration constraints shaped how much restructuring was needed to form a standardized final note (P1, P3, P4, P6, P8, P9, P12, P16, P18–20, P22, P23, P25, P28, P29). Clinicians described barriers when AI drafts did not fit EHRs’ problem-oriented structure that “saves under that particular diagnosis” (P4), requiring “copy and paste” steps (P4, P19). Additionally, clinicians deleted duplication when information was already documented elsewhere outside the AI smart elements (P8, P29). Others pointed to integration boundaries and chart access, including uncertainty about what the tool pulled beyond the conversation (P3, P24) and requests to the “ability to read the EHR” to support reusable history, continuity across visits, and real-time review (P1, P16, P18, P23).

### Clinician Expectations and Recommendations to Improve Ambient AI Draft Usability

We group recommendations by the scope of change and who would need to act on them, ranging from individual clinician behaviors and targeted system or configuration adjustments to broader workflow redesign and EHR integration. This framing highlights actionable informatics targets for individual clinicians, vendors, and health systems. Figure 1 provides an overview of the recurring challenges clinicians described and the improvement targets they recommended.

**Figure 1.**
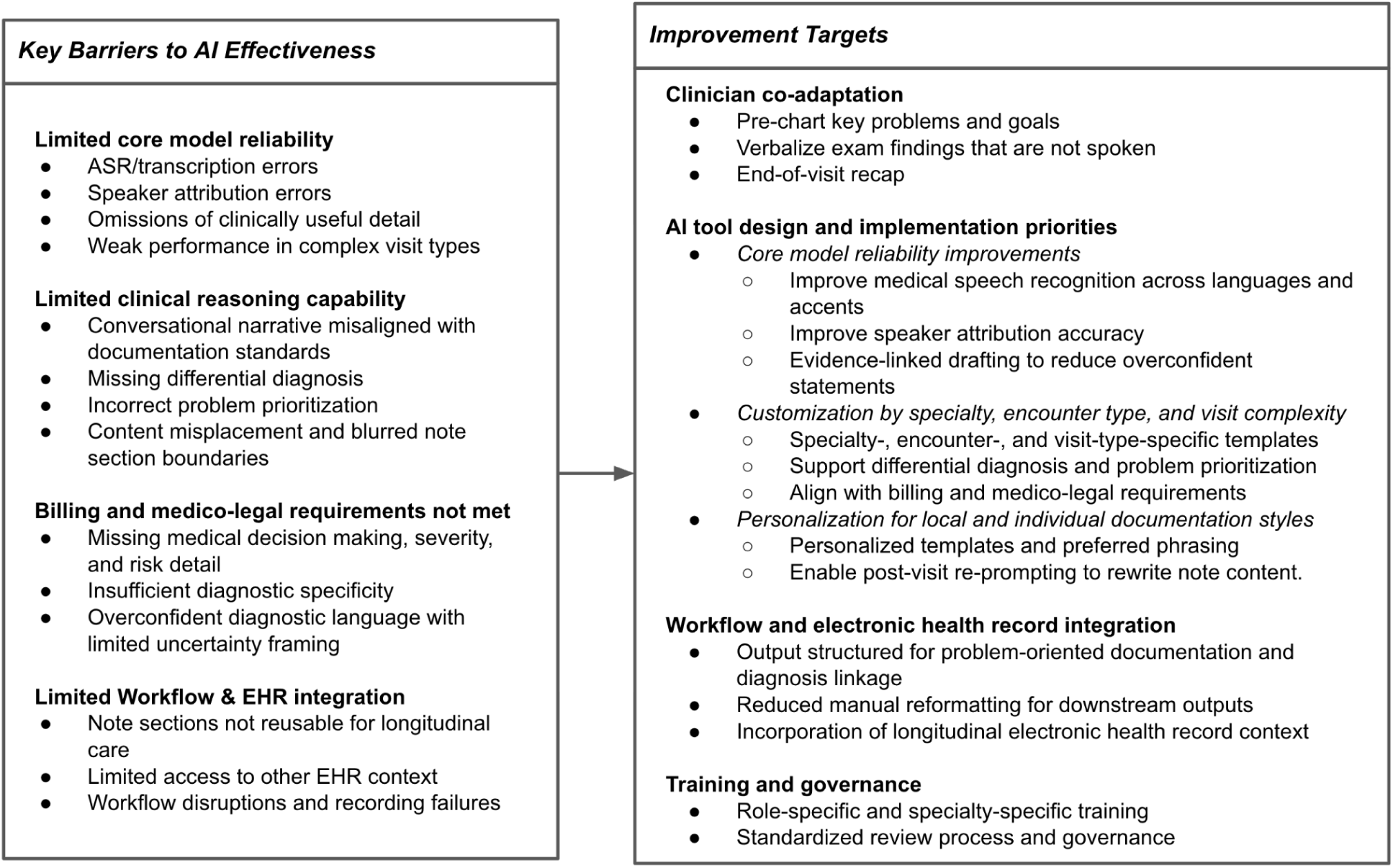
Challenges shaping ambient AI note editing and improvement targets

#### Precharting, in-visit verbalization, and post-visit strategies

To reduce editing load during note review, clinicians described adapting how they communicate so the ambient system can capture clinically relevant details more reliably (P3, P4, P6–14, P15–20, P26). A common practice was to verbalize information that is otherwise nonverbal, explicitly stating laterality and location when patients gesture (P3, P10, P11, P12, P13, P14, P17, P18, P19, P20). For example, “If a patient points, I’ll say out loud, ‘It looks like there’s a rash on your right arm,’ so AI can capture it” (P10). Others reported narrating exam maneuvers and findings in real time (“Okay, I’m doing a rectal exam… now I’m doing anoscopy…”) to populate the physical exam more completely (P6), or deliberately speaking standardized exam descriptors aloud (“regular rhythm,” “lungs are clear”) (P15). Outside the encounter, participants described pre-charting (P26, P9) and end-of-visit “wrap-up” strategies to reduce downstream edits, such as “summarizing the encounter at the end” (P6) or recapping problem-by-problem plans (“Okay, for the UTIs we’re going to do this…”) (P16).

#### Core AI model reliability, customization, and personalization for clinician-ready drafts

Clinicians’ vendor-facing recommendations clustered into three layers: improving core default behaviors, enabling specialty and encounter-type customization, and providing personalization controls. First, they described foundational model behaviors that should improve by default to reduce downstream edits. They emphasized fidelity to the encounter, preferring the system to “record what the clinician says” rather than produce “its own version” (P14). Several recommended accurate speaker attributions so the draft reflects who performed the assessment (P2, P5, P10, P15, P29). They also recommended conservative certainty as a default because “we tend to hedge more in medicine” (P2) and anticipated “fewer corrections…if hedging terminology is included by default” (P12), with certainty wording scaled to evidentiary strength (P26). In addition, participants highlighted gaps in medication and supplement handling (P17) and inconsistent recognition of the same drug across mentions (P18), motivating iterative updates of medication library (P21). Second, participants recommended specialty- and encounter-type customization, particularly for contexts where a generic draft does not fit, such as learner settings where “it has to be shorter,” with “a template for the AI to fit into an attestation” (P1), and specialties with “pretty standardized…how we were trained” documentation that may require different prompting to support the expected structure (P17). Third, clinicians wanted personalization functions, including the ability to “tweak it…more verbose, less verbose, what to include” and “give it more instruction” (P6), as well as re-writing controls such as “re-prompting ability like in ChatGPT” for targeted revisions (P1, P21, P30).

#### EHR integration to streamline workflow and reuse prior chart context

Clinicians recommended tighter EHR integration to reduce workflow friction and better support longitudinal care (P1, P4, P6, P8, P16–20, P22, P24–26, P28, P30). They suggested minimizing extra clicks and copy/paste for tasks such as after-visit summaries (“copy and paste…one paragraph”) (P8, P19). Participants also wanted AI outputs to sync with Epic structures (e.g., problem list and diagnosis-linked A&P) and reuse prior chart context—similar to dot phrases and structured history—to improve chronic disease management and longitudinal review (“a whole different experience”) (P16, P20, P22, P28). Additional integration expectations included drafting orders mentioned in conversation (“put that prescription in”), summarizing relevant chart review for complex histories, and improving specialty alignment so irrelevant cross-specialty content is omitted by default (P20, P24, P1, P19, P30). Finally, participants noted that more integrated capture workflows could reduce dependence on personal mobile devices and related interruptions (e.g., battery drain) (“phone died…charge during the visit”) (P1, P2, P18, P29).

#### Institutional Training and Governance to Reduce Uncertainty and Misuse

Clinicians emphasized that onboarding for ambient AI was often generic and recommended role-, setting-, and specialty-specific training, alongside clearer institutional guidance on when and how to obtain and document patient consent for ambient AI recording and documentation (P1, P2, P6, P24). They wanted practical instruction on “how to make it work,” noting that limited guidance can lead clinicians to rely on trial-and-error and may contribute to frustration or abandonment when workflows vary across settings and are not equally supported (P1).

Participants also highlighted broader education gaps in AI-assisted documentation that shape how clinicians evaluate and edit AI drafts (P2). From a governance perspective, they recommended standardizing consent processes, so the burden does not fall on individual clinicians each visit, and ensuring training covers routine workflows such as pre-charting and after-visit tasks (P6, P24).

## Discussion

Prior studies describe what clinicians edit in ambient AI notes. Our study is among the first to systematically characterize the underlying clinical, legal, and workflow rationales that drive those edits, translating edit behaviors into actionable design and governance. Clinicians whose documentation required more complex differential reasoning and were subject to greater billing and medicolegal scrutiny reported spending more effort correcting errors, restoring missing or misplaced clinical details, and rewriting content to align with specialty norms and evidentiary support. As a result, clinicians often either edit extensively or limit use to certain note sections they consider more reliable, which helps explain why adoption and perceived value vary by specialty and encounter type. [14]

Clinicians’ selective use across note sections often reflects systematic gaps between conversational summarization and the requirements of clinical documentation. [16] Accurate conversation capture does not guarantee clinically appropriate notes because documentation requires additional judgments about what is clinically relevant, where it belongs in the note, who the source is, and how strongly a statement should be made given available evidence and uncertainty. [17–19] These demands are more pronounced in subspecialty care, where notes require more longitudinal decision-making and specialty-specific differential reasoning while also meeting billing, legal, and institutional expectations. [12,13,20] In this setting, recurring limitations such as transcription errors and omissions of key clinical details can cascade into substantial downstream editing. The problem is further intensified when drafts are overly long, fail to distinguish clinically relevant content, or do not align with EHR workflows. [21]

Improving ambient AI tools will require coordinated, multi-level efforts. At the foundational model level, systems should prioritize robust transcription and accurate speaker attribution. [22] When they fail, downstream content becomes harder to trust and more time-consuming to review. Drafts should also default to safer clinical language by tying statements to evidence and using conservative certainty when the basis is unclear. [23–25] Beyond baseline reliability, ambient drafting should better reflect specialty and encounter type needs through configurable templates and encounter presets, such as new visits versus follow-up visits, chronic disease management versus procedures, and visits involving learner supervision versus independent practice. [26,27] However, we do not know how comprehensive or deliberate clinicians are in verbalizing the information they intend to include in the note; ambient drafts can only draw from what is stated in the encounter transcript. When key procedural rationale or context is not explicitly articulated, drafts may default to hedging or inference rather than documenting clear justification. Future work should evaluate “speak-to-document” practices and test whether specialty- and visit-type templates can cue more structured, note-ready clinical language at the point of care. Furthermore, ambient AI should be implemented as an EHR-integrated workflow rather than a standalone note generator. Access to the patient history, medication list, and recent notes are essential for longitudinal care and can prevent redundant information that clinicians routinely delete in drafts. [28,29] Finally, because many edits reflect personal preferences, tools should support clinician-level personalization through reusable templates, and controlled learning from prior edits, paired with pre-prompting or re-prompting functions to reduce repetitive rewriting and corrections. [30]

In addition, institutions should provide thorough implementation support through specialty-, role-, and workflow-specific onboarding [31,32], alongside clear governance guidance and a standardized review checklist specifying how and what clinicians should verify. [33] Clinicians may also benefit from actively adopting “AI-aware” communication and review behaviors to improve draft quality and support human–AI collaboration.

Our study has several limitations. These insights reflect outpatient workflows and a self-selected group of participating clinicians and may underrepresent non-adopters and settings such as inpatient or emergency care where documentation practices and operational constraints differ.

Findings are also limited to a single institution and a single ambient AI vendor, which may affect generalizability. Despite these limitations, the results suggest that sustained reductions in editing burden will require closer alignment between ambient AI outputs and real-world documentation workflows. Such alignment may help shift ambient AI from producing generic summaries that require substantial rewriting to generating drafts that are faster to review, and more consistently usable across specialties.

## Conclusion

When editing the ambient AI documentation drafts, clinicians aim to safeguard factual accuracy, preserve specialty-appropriate reasoning and terminology, and ensure documentation meets billing, compliance, and medico-legal expectations. Based on these findings, we recommend improving core draft reliability by increasing transcription and speaker attribution accuracy and using default language that appropriately reflects clinical uncertainty. We also recommend adaptable specialty- and encounter-specific templates, stronger EHR integration to incorporate longitudinal context, and institutional training and governance to support consistent, safe AI-assisted documentation practices.

## Data Availability

De-identified materials may be made available to researchers upon reasonable request to the corresponding author and approval by the relevant institutional authorities.

## ACKNOWLEDGEMENT

We thank the UCI Health clinicians who participated in this study and shared their time and insights. We also appreciate the institutional support from UCI Health that enabled recruitment and data collection.

## CONTRIBUTORSHIP STATEMENT

Yawen Guo (YG) led the study and contributed to study conception and design, interview protocol development, data collection, qualitative analysis and interpretation, and manuscript drafting. Di Hu (DH) contributed to interview protocol development, qualitative coding and analysis. Ziqi Yang (ZY) contributed to qualitative coding and analysis. Steven Tam (ST), Emilie Chow (EC), Danielle Perret (DPe), and Deepti Pandita (DPa) provided clinical input and supported interview piloting and participant recruitment. Kai Zheng (KZ) provided overall supervision and guidance throughout the project. All authors critically reviewed and revised the manuscript for important intellectual content and approved the final version.

